# Impact of neuromodulation on post-stroke aphasia: a multimodal randomized controlled study

**DOI:** 10.1101/2023.02.12.23285828

**Authors:** Zhong Sheng Zheng, Kevin Xing-Long Wang, Henry Millan, Sharon Lee, Melissa Howard, Aaron Rothbart, Joel Frohlich, Emily Rosario, Caroline Schnakers

## Abstract

**Background:** Transcranial direct current stimulation (tDCS) combined with speech and language therapy (SLT) may increase the effectiveness of recovery in post-stroke aphasia. However, inconsistent responses have been observed, in part due to small sample sizes, limited comprehensive assessments, and poor mechanistic understanding of intervention related recovery.

**Methods:** Using a double-blind, randomized, sham-controlled design, we investigated the efficacy of anodal Broca’s tDCS combined with SLT over five 20-minute daily sessions in 45 chronic stroke patients. EEG and DTI were used to explore treatment-mediated neuroplastic mechanisms. The primary outcome measures were linguistic skills evaluated by the Western Aphasia Battery-Revised before and after the intervention.

**Results:** Compared to sham (SLT + placebo), tDCS patients improved significantly more in aphasia quotient, auditory verbal comprehension, and spontaneous speech. While tDCS improved both expressive and receptive domains, sham only improved expressive language. EEG showed recovery in both groups to rely predominantly on the contralesional side, particularly the right middle temporal area (T4). While tDCS induced recovery correlated with changes in the faster frequencies (e.g., alpha, beta), sham mediated recovery correlated with changes in the slower frequencies (e.g., theta, delta). Furthermore, reduced beta coherence between T3 and T4 was associated with repetition gains specific to tDCS. Furthermore, improved spontaneous speech in tDCS was associated with decreased mean diffusivity in superior cerebellar peduncle. Given this region’s connectivity with contralateral cortical regions, this finding extends and aligns with the EEG signatures of neuroplasticity in right-lateralized cortical regions, highlighting the role of cerebro-cerebellar connections in language recovery.

**Conclusions:** Our findings support the use of anodal Broca’s tDCS for enhancing both expressive and receptive language domains in chronic aphasia. To the best of our knowledge, this study represents the first multimodal neuroimaging (EEG, DTI) study to uncover mechanistic differences between tDCS and behavioral mediated aphasia recovery, and the first to identify a cerebellar white matter marker of language recovery following tDCS.

**Clinical Trial Registration:** http://www.clinicaltrials.gov [NCT03699930]

## Introduction

The difficulty of using language to communicate and express one’s thoughts directly challenges the centrality of human existence; relaying our internal world to others with high fidelity is a huge factor in what separates us from any other species. As stroke remains the leading cause of long-term disability in the United States, it is estimated that up to 38% of stroke patients suffer from aphasia,^1^ a language disorder severely impacting daily functioning and quality of life.^2^ Stroke survivors with aphasia also show greater morbidity and mortality than patients without aphasia. As such, it should come as no surprise that the recovery of language abilities is a top priority amongst stroke survivors.^3^

Transcranial direct current stimulation (tDCS) has emerged as a popular adjunct therapy to traditional speech and language therapy (SLT) in post-stroke aphasics to facilitate language recovery by enhancing beneficial and suppressing maladaptive plasticity. Increasing evidence suggests that SLT combined with tDCS is more effective than SLT alone.^4–8^ While promising, studies investigating the efficacy of tDCS have produced variable responses, likely in part due to small sample sizes, limited comprehensive assessments (often limited to naming tasks), and poor mechanistic understanding of tDCS-mediated recovery. tDCS studies that employ double-blind randomized controlled trials using a multimodal approach in large samples remain sparse in chronic post-stroke aphasia.

Resting-state EEG (rsEEG) represents an ideal technique to assess objectively the effect of neurorehabilitation treatment in patients with acquired brain injury since it is low cost, portable, and widely available in hospital settings. Previous studies have mostly shown a slower brain activity following stroke in acute and subacute patients as compared to controls (e.g., increase in delta and theta waves and decrease of faster frequencies such as alpha and beta waves).^9^ Such characteristics were related to behavioral outcome and were suggested to change across time, with slower frequencies decreasing and faster frequencies increasing over time.^10–13^ Additionally, functional disconnection between key areas (such as Broca’s and Wernicke’s) within the left hemisphere and with the contralesional hemisphere has also been reported following stroke.^14,15^ To our knowledge, only two studies^16,17^ have assessed electrophysiological changes after tDCS (of Broca’s area, the primary motor cortex, and the posterior perisylvian region; all on the left hemisphere) in a small sample size (*n =* 12) of patients with post-stroke aphasia. Therefore, there remains a critical need for better understanding electrical brain changes following such treatment in this population.

In addition to examining functional changes by EEG, which is limited to the cortex, we also used diffusion tensor imaging (DTI) to study white matter microstructural changes after intervention. Given its sensitivity to short-term interventions and its availability in most hospitals, DTI represents another feasible clinical and research tool for studying disease recovery. The two most common DTI parameters, fractional Anisotropy (FA) and mean diffusivity (MD), can be used to infer white matter organization and integrity.^18^ Studies that have used DTI to study mechanisms underlying disease and recovery processes in post-stroke aphasia have mostly examined the arcuate fasciculus, a white matter tract connecting Broca’s with Wernicke’s area, or other supratentorial white matter regions.^19^ However, delineating supratentorial white matter tracts can be labor intensive and challenging given the anatomical distortion from large stroke lesions (typically affecting supratentorial areas in aphasia), rendering this approach not automatable and infeasible in a clinical setting. An alternative strategy could be to explore more intact regions that are indirectly connected with cortical language nodes, such as white matter regions below the tentorium (i.e., infratentorium), which are relatively spared in post-stroke aphasics, as the cerebellum is structurally connected with contralateral language networks via the brainstem.^20^ We believe this type of investigation has yet to be carried out before in post-stroke aphasia.

In this double-blind randomized controlled study, we aim 1) to evaluate the efficacy of tDCS coupled with SLT in a large sample of chronic post-stroke aphasics (*n* = 45) using the Western Aphasia Battery-Revised and 2) to understand intervention related neural changes using EEG and DTI. By combining these two methods, we can gain a more complete understanding of both functional and structural changes that occur as a result of different treatments. This is the first study to incorporate a broad assessment of language domains coupled with EEG and DTI in the context of tDCS intervention for post-stroke aphasia.

## Materials and methods

### Experimental design

Following a double-blind randomized controlled design, patients were randomly assigned to receive either the active anodal (1mA) left inferior frontal (Broca’s area) or sham stimulation, concurrent with 20 minutes of speech and language therapy by a trained speech language pathologist over five consecutive days. A random block allocation sequence was computer-generated to assign participants to either tDCS or sham in a 1:1 ratio. The sequence consisted of a series of codes (i.e.,6 digits). One code was attributed per patient following the sequence order. Each code was provided by the principal investigator (PI) to the experimenter on the day of the patient’s first tDCS session. This code was encoded in the tDCS device before starting each session, blinding both the experimenter and the participant. Only the PI was aware of the allocation and was not involved in data collection or interaction with participants. The PI did not discuss the allocation until data analysis. Data analyses were not blinded. Behavioral, EEG, and DTI data were collected within one week before and after intervention. Moreover, secondary behavioral outcome measures were additionally collected at 3-month follow-up.

### Transcranial direct current stimulation

Using the Soterix Medical 1×1 tDCS system (Soterix Medical Inc., New York, NY), electrodes were placed inside saline-soaked sponges (5 cm x 7 cm) to deliver a current of 1 mA over the scalp. Based on the International 10-20 system for EEG electrode placement, the anode was placed over F7 corresponding to Broca’s area; the cathode was placed over Fp2, the right frontopolar cortex.^7^ For the active condition, stimulation included an initial ramp-up period of 30 seconds to the 1mA intensity and was maintained for 20 minutes before ramping down to 0 mA over 30 seconds toward the end. For the sham condition, the initial 30 second ramp-up was immediately followed by a 30 second ramp-down, with intensity maintained at 0 mA during the session. Both stimulation conditions were paired with 20 minutes of speech and language therapy. All participants underwent five consecutive days of stimulation.

### Speech and language therapy

Based on results from standardized assessment measures, the 20-minute standard of care therapy session incorporated techniques from Melodic Intonation Therapy (MIT), language stimulation approach, modified response elaboration training, Semantic Feature Analysis Treatment, Verb Network Strengthening Treatment (VNeST), Sound Production Treatment, Integral Stimulation, and Word Retrieval Cuing Strategies (e.g., phonological and semantic cuing). Auditory comprehension tasks were indirectly embedded into therapy, especially for participants with severe auditory comprehension deficits. Therapy targeted areas that would be most impactful on functional communication specific to each participant.

### Participants

Patient inclusion criteria consisted of aphasia (diagnosed by a neurologist or speech and language pathologist) due to ischemic or hemorrhagic stroke, at least 6 months post stroke, over 18 years old, and English speaking. All patients suffered a left hemisphere stroke. Patients were excluded if they had other neurologic conditions other than stroke. Data were collected between 2018 and 2020. This study can be broken down into three components: behavioral, EEG, and DTI. Refer to Fig. 1 and Table 1 for the breakdown of participants and their demographic and clinical information. Forty-five patients (20 tDCS; 25 sham) with chronic post-stroke aphasia were included in the behavioral component. A subset of them contained complete neuroimaging data, with 40 patients (19 tDCS; 21 sham) in the EEG and 33 patients (16 tDCS; 17 sham) in the DTI analyses. On the basis of a type I error rate of 5% and a power of 80%, a total sample size of 44 was determined for a two-way mixed ANOVA to reach a 5-point difference considered clinically meaningful on the Western Aphasia Battery-Revised (WAB-R) score.^21,22^ This study was approved by the Institutional Review Board at Casa Colina Hospital and Centers for Healthcare. All participants provided written informed consent prior to participation (Registered on clinicaltrials.gov as NCT03699930).

**Figure 1.**
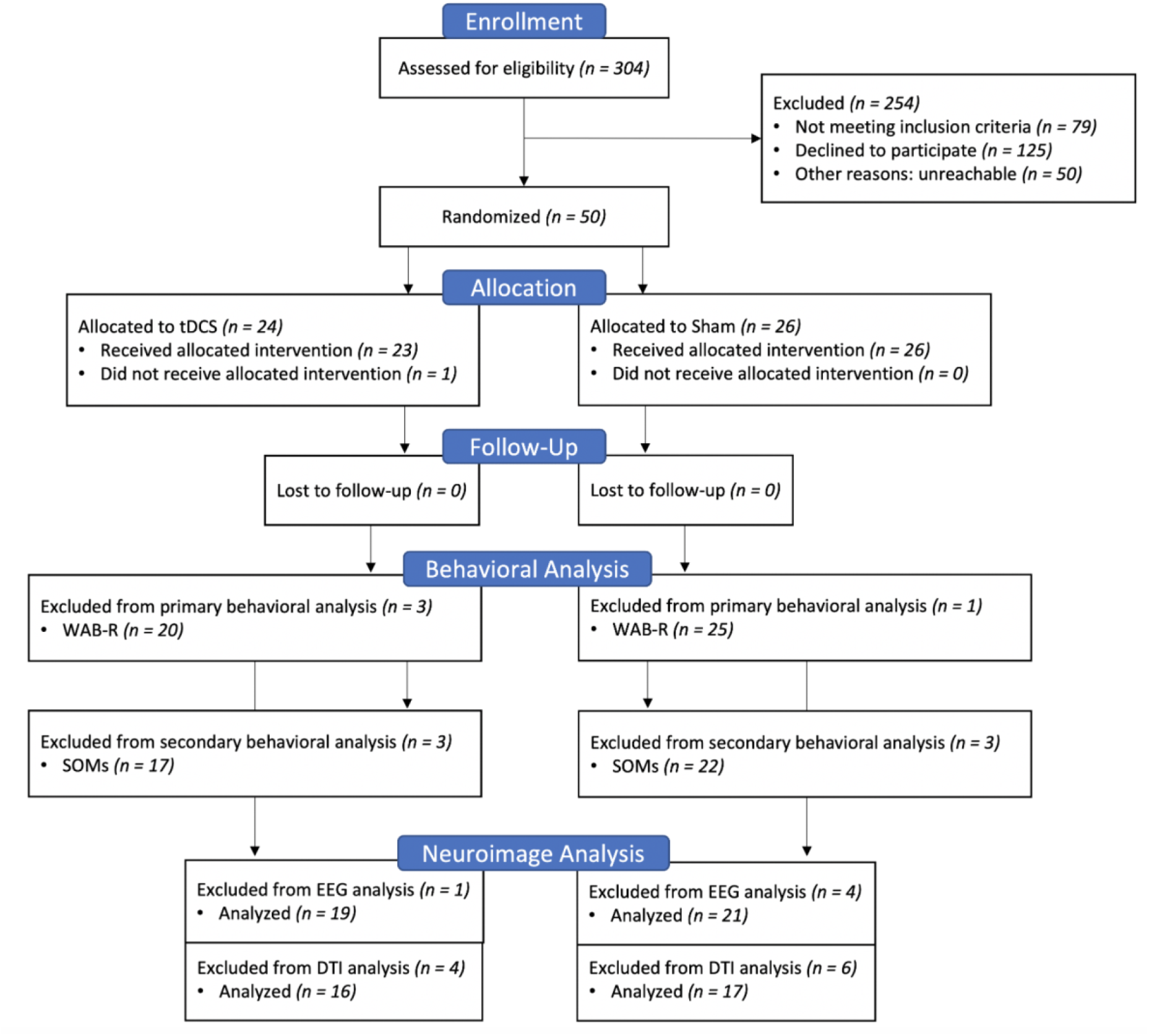
Consolidated Standards of Reporting Trials (CONSORT) Flow Diagram.

**Table 1.** Patient Demographic and Clinical Information.

| Variable | Behavioral |  | EEG |  | DTI |  |
| --- | --- | --- | --- | --- | --- | --- |
|  | tDCS | Sham | tDCS | Sham | tDCS | Sham |
| N | 20 | 25 | 19 | 21 | 16 | 17 |
| Age (years) | 55 ± 15.6 | 64 ± 11.1 | 57 ± 13.7 | 64 ± 12 | 55 ± 15.1 | 63 ± 11.5 |
| TSI (months) | 49.3 ± 31.6 | 87.7 ± 74.8 | 49.9 ± 32.4 | 85.4 ± 77.5 | 48.1 ± 29.8 | 76 ± 69.4 |
| Baseline AQ | 60.4 ± 26.9 | 60.3 ± 28.5 | 60 ± 27.4 | 62 ± 29.3 | 57 ± 25.6 | 62 ± 29.6 |
| Sex (%) |  |  |  |  |  |  |
| Male | 75 | 80 | 78.9 | 81 | 81.3 | 76.5 |
| Female | 25 | 20 | 21.1 | 19 | 18.8 | 23.5 |
| Aphasia Type (%) |  |  |  |  |  |  |
| Fluent | 55 | 52 | 52.6 | 57.1 | 43.8 | 58.8 |
| Non-Fluent | 45 | 48 | 47.4 | 42.9 | 56.3 | 41.2 |
| Stroke Type (%) |  |  |  |  |  |  |
| Ischemic | 75 | 76 | 78.9 | 76.2 | 75 | 70.6 |
| Hemorrhagic | 20 | 20 | 21.1 | 19 | 18.8 | 23.5 |
| Both | 5 | 4 | 0 | 4.8 | 6.3 | 5.9 |

### Behavioral outcome measures

Behavioral assessments were conducted by a speech-language pathologist in English. The WAB-R was used to assess aphasia severity (aphasia quotient [AQ]) and consisted of four subscales addressing spontaneous speech (SS), name and word finding (NWF), repetition (REP), and auditory verbal comprehension (AVC). The primary outcome was improvement in language and communication as measured by WAB-R scores. The secondary outcome measures (SOMs) were self-reported and included Communication Outcomes after Stroke (COAST), Carer COAST, Stroke and Aphasia Quality of Life Scale-39 (SAQOL-39), as well as Patient-Reported Outcomes Measurement Information System (PROMIS). All outcome measures were assessed before and after intervention, with an additional 3-month follow-up for secondary behavioral outcome measures only.

### EEG recording

Electrophysiological data of each subject was recorded using the wireless B-Alert X-24 EEG System (B-Alert wireless EEG system; advancedbrainmonitoring.com). EEG data were acquired at the Casa Colina research institute in a Faraday room within one week before and after intervention. A 24-electrode skull cap was connected to a wireless portable digital EEG amplifier. Fp1 and Fp2 were used to monitor eye artifacts. A mastoid reference was applied. EEG-data were recorded on a laptop computer. The impedances were kept below 40 kΩ. A sampling rate of 250 Hz was used. EEG recordings were performed while the participants were in a wakeful state with eyes open, sitting in a setting with minimal ambient noise. Patients were instructed to relax and look at a fixed object in the room (i.e., doorknob). Duration of recording was around 10 minutes.

### MRI acquisition

MRI was acquired on a Siemens Magnetom Verio scanner (3 Tesla) within one week before and after intervention. The following images were collected and used for analysis: 3D T1-weighted MPRAGE (TR = 2300 ms; TE = 2 ms; flip angle = 9°; FOV = 230 mm; slice thickness = 1 mm with no gap; number of slices = 160; matrix size = 224 × 224), diffusion weighted imaging (TR = 10900 ms; TE = 95 ms; slice thickness = 2 mm with no gap; number of slices = 82; matrix size = 122 × 122; 64 diffusion directions [*b* = 1000 s/mm^2^] with five B0’s).

### Data analysis

For each dataset (behavioral, EEG, DTI), we first determined whether or not the two groups significantly differed in age, sex, baseline AQ (i.e., aphasia severity), type of stroke (hemorrhagic, ischemic), aphasia type/fluency (fluent, non-fluent), and time since injury (TSI), using independent t-tests and X^2^ tests, as appropriate. Variables reflecting significant differences between groups were considered as covariates. Statistical tests were two-tailed.

#### Behavioral

Using SPSS, two-way mixed ANOVAs, with group (tDCS, sham) as the between-subjects factor and time (pre, post) as the within-subjects factor, were carried out for each WAB-R score (AQ, SS, AVC, NWF, and REP) while covarying out age and TSI to determine whether the change from pre to post intervention is different between tDCS and sham. Partial eta squared (ηp^2^) was used for measuring effect sizes.

#### EEG

##### Preprocessing

EEG data were artifact-reduced in EEGLAB ^23^ using a combination of manual cleaning and independent component analysis (ICA). Manual data cleaning entailed marking and excising sections of EEG recordings with gross physiological artifacts. Next, we performed ICA using EEGLAB’s runica function, which implements the Infomax ICA algorithm by,^24^ and removed ICs corresponding to stereotyped physiological artifacts (e.g., eye movements, muscle activity, or ballistocardiogram).

##### EEG analysis

Following ICA cleaning, all data were re-referenced to average. We then computed 1 – 25 Hz spectral power in all channels using 2 s Hamming windows with 75% overlap (frequency bin width: 1 Hz). Next, we computed the imaginary coherence, a measure of coherence (i.e., phase correlation)^25^ that is robust to spurious results due to volume conduction artifacts. Specifically, we computed frontotemporal functional connectivity as the imaginary coherence (IC) between two frontal channels (F7, F8) and two temporal channels (T3, T4) using the Brainstorm toolbox^26^ with the function bst_cohn and the coherence option ‘icohere2019’; as with spectral power, we used 2 s windows with 75% overlap. In both analyses (spectral power and imaginary coherence), windows that contained temporal discontinuities where noisy data segments had been removed were excluded to avoid Gibbs phenomena and similar artifacts. Spectral power at each frequency bin was examined as relative power (i.e., normalized by the total integrated 1 – 25 Hz power), and all power values were log_10_ transformed prior to analysis. The frontotemporal channels of interest were chosen based on proximity to the stimulation location (F7/left inferior frontal/putative Broca’s area) and included the closest temporal channel (T3/left middle temporal/near Wernicke’s area) and the right hemisphere homologs (F8, T4).

##### Statistical analysis

To better understand the neural mechanisms underlying intervention related language improvement, we correlated pre/post changes in relative power and coherence with pre/post changes in WAB-R scores for tDCS (tDCS + speech therapy) and sham (placebo + speech therapy) separately using Spearman’s rank, while covarying out age and TSI. To correct for multiple comparisons, we applied the Benjamini-Hochberg method with a false discovery rate (FDR) of 0.1. This FDR threshold was chosen to balance the cost of false positives and negatives in light of the exploratory nature of this investigation. Finally, Fisher’s r-to-z transformation was used to statistically compare the correlation coefficients between tDCS and sham upon finding a significant correlation in either group.

#### MRI

##### Preprocessing and lesion identification

Using FSL tools (http://www.fmrib.ox.ac.uk/fsl), T1-weighted images first underwent bias-field correction. Then, brain extraction was performed with optiBET (Optimized Brain Extraction for Pathological Brains).^27^ For stroke lesion tracings, we employed an automated segmentation algorithm, LINDA (Lesion Identification with Neighborhood Data Analysis), by supplying patients’ T1-weighted images.^28^ Lesion masks resulting from LINDA were manually edited as needed. Individual T1-weighted lesion masks were transformed into standard space (MNI 1mm) using Advanced Normalization Tools (ANTs).^29^ Once in standard space, all the lesion masks were binarized and combined to create a group lesion map. For diffusion-weighted data preprocessing, DTIPrep^30^ was used to automatically identify and remove volumes with substantial artifacts as well as to correct for eddy currents and head motion with affine registration to the average *b* = 0 reference volume. BET (Brain Extraction Tool) was used to skullstrip the *b* = 0 image.^31^

##### DTI analysis

Diffusion tensors were calculated for each voxel of the brain using the diffusion toolbox in FSL, and FA and MD maps were subsequently computed. Implementing a region of interest (ROI) approach, we used labels from the Johns Hopkins white matter atlas (http://cmrm.med.jhmi.edu). However, due to the heterogeneity and extensiveness of the stroke lesions, which mainly affected regions above the tentorium, preventing accurate delineation of the ROIs, we opted to only investigate regions below the tentorium, which were relatively intact across all patients. Eleven ROIs were selected (Supplementary Fig. 1): medial lemniscus (ML), pontine crossing tract (PCT), middle cerebellar peduncle (MCP), bilateral cerebral peduncle (CP), bilateral corticospinal tract (CST), bilateral superior cerebellar peduncle (SCP), and bilateral inferior cerebellar peduncle (ICP). By applying nonlinear registration between the patients’ diffusion and standard space (FMRIB58_FA), these standard ROIs were transformed into each patient’s diffusion space. Prior to data analysis, all ROIs were visually examined for accuracy and manually corrected as needed. To further refine the ROIs and ensure that only white matter voxels are included, a whole-brain white matter segmentation mask was created for each patient to intersect with the ROIs. The average fractional anisotropy (FA) and mean diffusivity (MD) values of each ROI were calculated for each patient in diffusion space.

##### Statistical analysis

Covariate-adjusted Spearman’s rank correlation was carried out between the change (post – pre) in DTI metrics (FA and MD) and change in the WAB-R scores. Again, Benjamini-Hochberg method (FDR = 0.1) was used for multiple comparisons correction. Fisher’s r-to-z transformation was employed for direct comparisons of correlation coefficients between groups.

## Results

The group lesion overlap map indicated damage mostly involving the left perisylvian regions, with the highest lesion overlap seen in the left superior longitudinal fasciculus (Fig. 2). Age and TSI were found to be significantly different between the tDCS and sham group; therefore, age and TSI were included as covariates in all subsequent analyses. No significant adverse events were reported.

**Figure 2.**
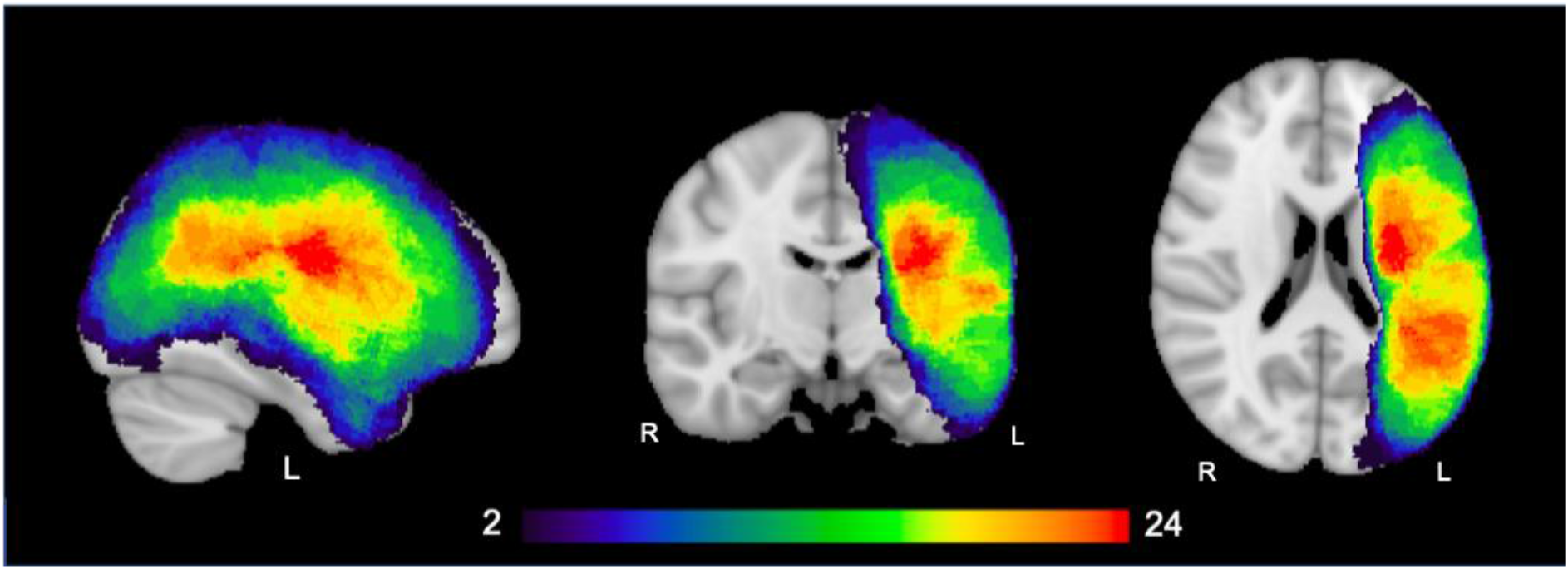
Group Lesion Overlap. The group lesion map shows areas of stroke damage with at least two patients overlapping. The left perisylvian regions were the hotspots of damage, with the highest lesion overlap observed in left superior longitudinal fasciculus. Color bar indicates the number of patients overlapping.

### Behavioral

Forty-five patients (20 tDCS; 25 sham) with chronic post-stroke aphasia were included in the analyses of the primary outcome measures and 39 patients (17 tDCS; 22 sham) in the analyses of the secondary outcome measures.

#### WAB-R scores

Group x Time interactions (Fig. 3) were significant for AQ (*F*(1, 41) = 6.415, *P* = 0.015, ηp^2^ = 0.14) and AVC (*F*(1, 41) = 6.444, *P* = 0.015, ηp^2^ = 0.14), and marginally significant for SS (*F*(1, 41) = 4.013, *P* = 0.052, ηp^2^ = 0.09). Post-hoc comparisons with Bonferroni correction revealed that tDCS exhibited greater improvement in AQ, SS, and AVC, as compared to sham. Moreover, both groups improved significantly after intervention for all the WAB-R scores, except for AVC and REP (trending significance at *P* = 0.058) in the sham group.

**Figure 3.**
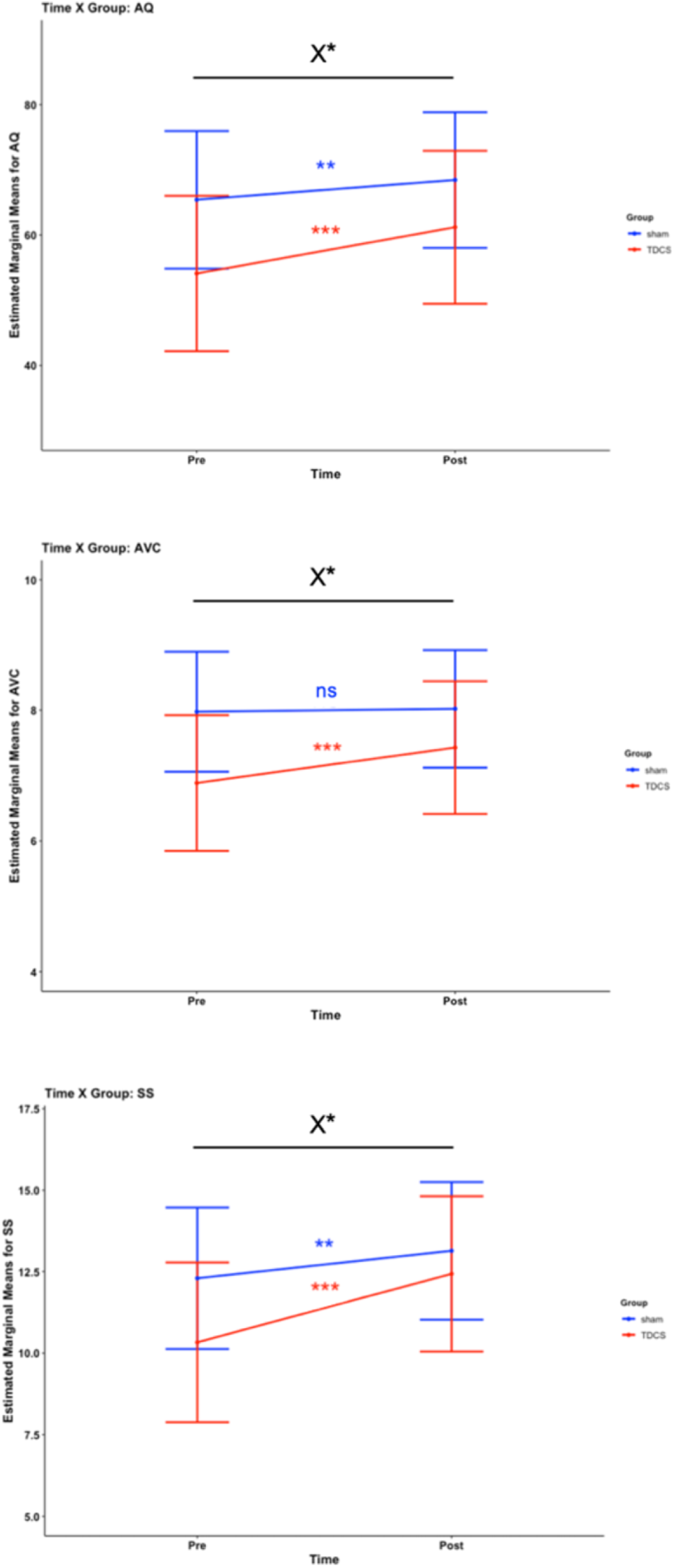
Behavioral Interaction Plots. Black line with X denotes an interaction effect. Blue indicates a significant difference between the pre- and post-scores for sham, whereas red indicates a significant difference between the pre- and post-scores for tDCS. Covariates appearing in the model are evaluated at the following values: TSI = 2150.56 days; Age = 60.07 years. Error bars: 95% CI. ns = non-significant; * = *P* ≤ 0.05; ** = *P* ≤ 0.01; *** = *P* ≤ 0.001.

#### Secondary outcome measures

No significant Group x Time interaction was found for any of the secondary outcome measures.

### EEG

#### Relative Power

Partial correlations (adjusting for TSI and age) using Spearman’s Rank identified different patterns of EEG correlation with WAB-R scores between tDCS and sham (Fig. 4). Reporting only the results surviving multiple comparisons correction (see Supplementary Table 1 for results that did not pass multiple comparisons correction), we found that, for tDCS, change in relative alpha power at T4 positively correlated with change in AQ (*r*_*s*_(15) = 0.664, *P* = 0.004) and SS (*r*_*s*_(15) = 0.705, *P* = 0.002). For sham, we found a negative correlation between change in relative delta power at T4 and change in AQ (*r*_*s*_(17) = -0.615, *P* = 0.005) and a positive correlation between change in relative delta power at F8 and change in REP (*r*_*s*_(17) = 0.628, *P* = 0.004). Directly comparing these correlation coefficients between tDCS and sham revealed a significant group difference for F8 – delta with repetition (z = 2.422, *P* = 0.015).

**Figure 4.**
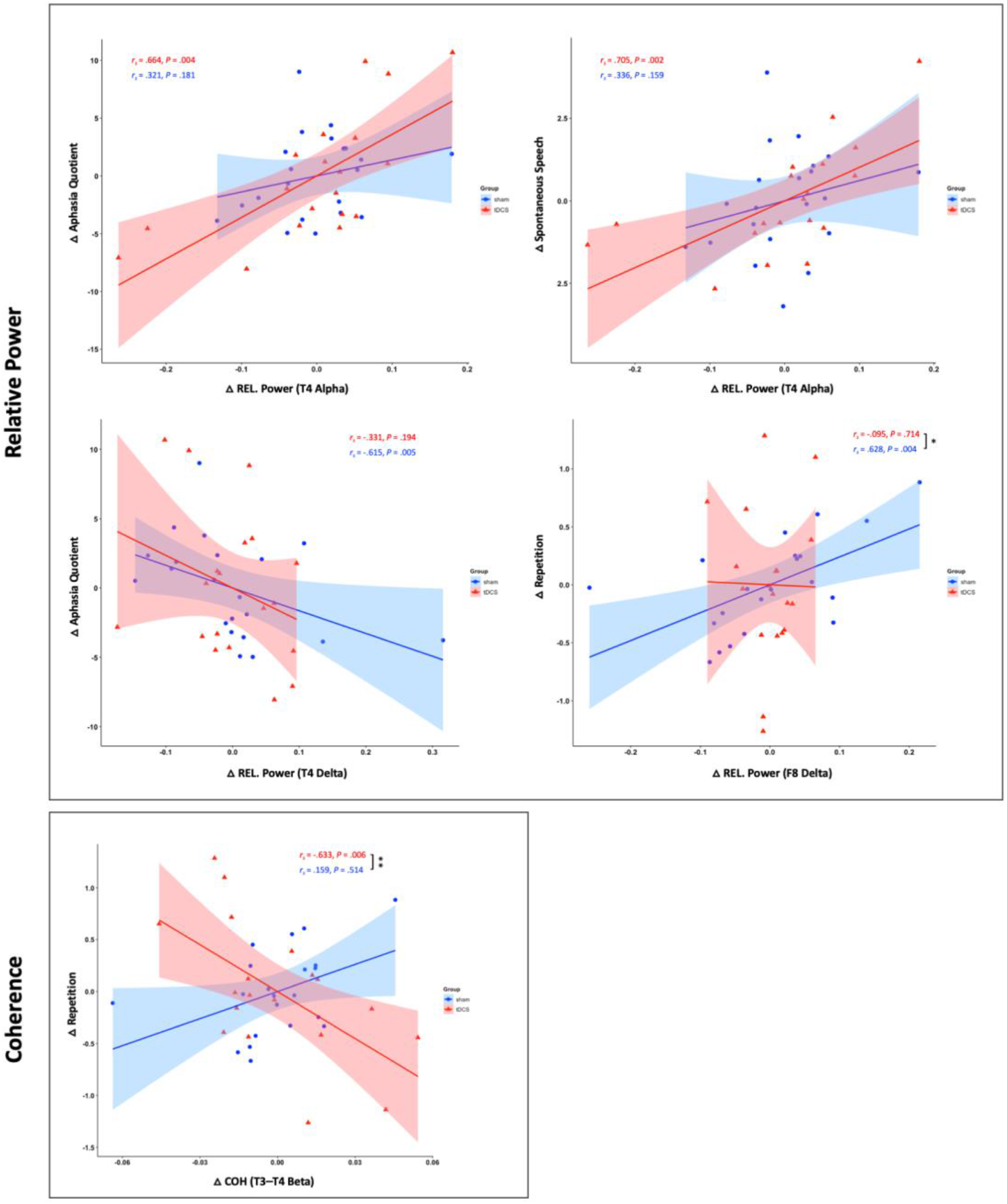
EEG and Behavioral Correlations. Partial correlations (controlling for age and TSI) between change (post – pre; Δ) in WAB-R scores and change in EEG parameters (relative power [REL Power], coherence [COH]) are displayed. Error bars reflect 95% confidence interval. Astericks (* = *P* ≤ 0.05; ** = *P* ≤ 0.01) represent significant correlation differences between tDCS and sham.

#### Imaginary Coherence

For tDCS, a negative partial correlation between the change in T3-T4 beta coherence and change in repetition score (*r*_*s*_(15) = -0.633, *P* = 0.006) (Fig. 4) was identified to be significantly different (z = 2.64, *P* = 0.008) from sham’s partial correlation.

### DTI

After multiple comparisons correction, we found that change in MD of the left superior cerebellar peduncle negatively correlated with change in spontaneous speech for the tDCS group (*r*_*s*_(12) = -0.777, *P* = 0.001), but not sham (Fig. 5). Refer to Supplementary Table 2 for both uncorrected and corrected findings. Comparing the groups’ correlation coefficients revealed a statistical difference (z = 3.120, *P* = 0.002).

**Figure 5.**
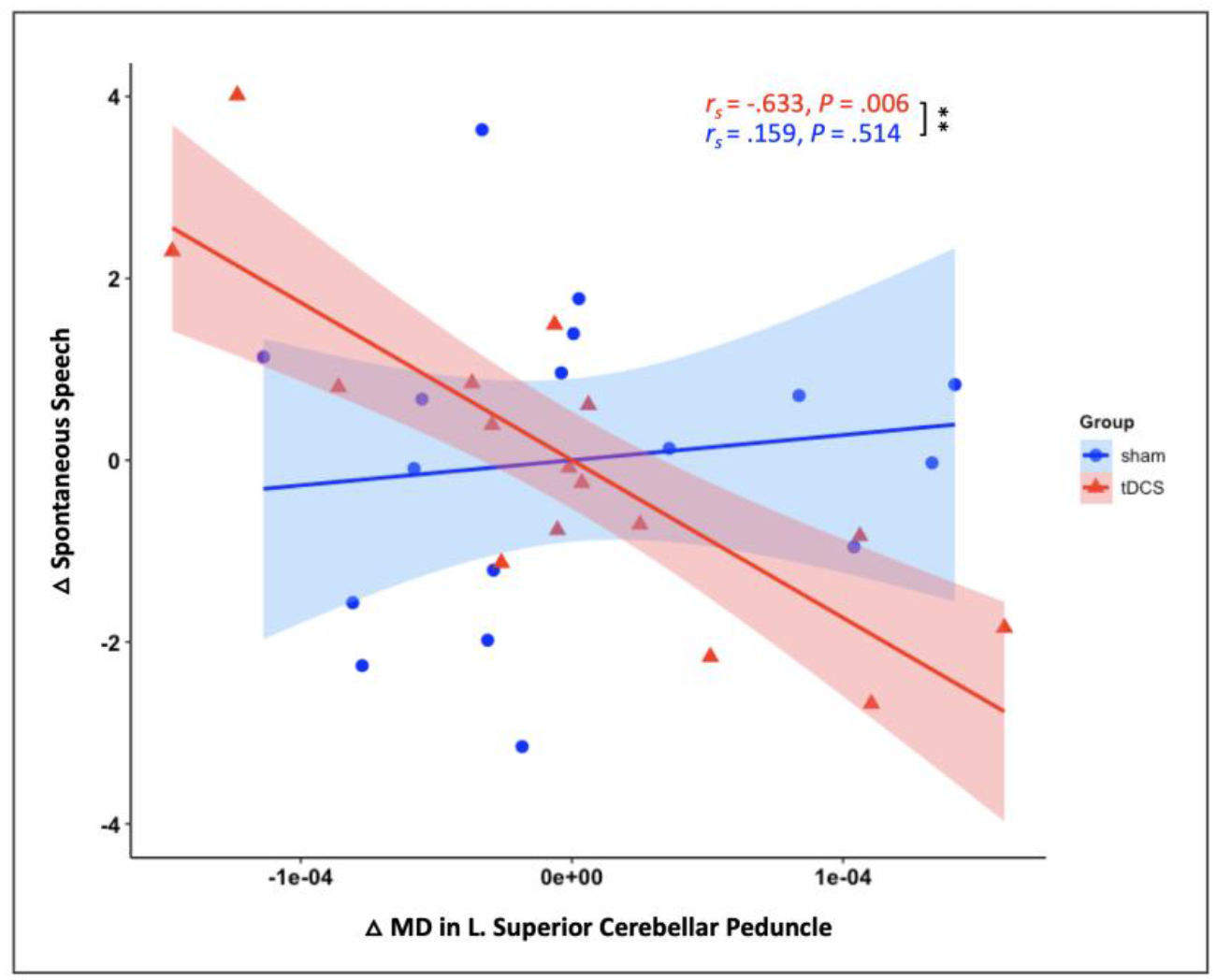
DTI and Behavioral Correlations. Partial correlations (controlling for age and TSI) between change (post – pre; Δ) in WAB-R scores and change in DTI parameter (mean diffusivitity [MD]) are displayed. Error bars reflect 95% confidence interval. Astericks (* = *P* ≤ 0.05; ** = *P* ≤ 0.01) represent significant correlation differences between tDCS and sham.

## Discussion

### Behavioral

Our results corroborate prior work supporting the use of anodal tDCS over the left inferior frontal gyrus (IFG; Broca’s area) as an adjunct therapy for chronic aphasia. Previous studies using anodal Broca’s tDCS have largely shown improvement in expressive language abilities, such as repetition,^32^ naming,^7,33,34^ and spontaneous speech.^35,36^ Due to the primary focus on language production in studies involving Broca’s area, the multi-faceted language effects of stimulating this brain region were not fully elucidated, as receptive language skills were not adequately evaluated in these studies. By evaluating both expressive and receptive language abilities using the WAB-R, we found that Broca’s tDCS not only improved language production (e.g., spontaneous speech) but also comprehension (e.g., auditory verbal comprehension). More recent studies also uncovered the effects of anodal Broca’s tDCS in enhancing sentence comprehension.^37,38^ Indeed, Broca’s area is known to mediate language production but can also contribute to comprehension.^39^ It is worth noting that while the WAB-R assesses a range of different language faculties, it may not be sensitive enough to detect small improvements. This may be the case for repetition performance, which showed a non-significant trend of tDCS patients improving more than sham (*P* = 0.1).

Finally, there were no significant differences between groups (tDCS vs. sham) in the self-reported (secondary) outcome measures across the three timepoints (pre, post, and 3-month follow-up) even though we found objective differences in the WAB-R assessments, suggesting the potential influence of a placebo effect and/or unreliability of the subjective reports, highlighting the importance of including objective measures in such protocol.

Unfortunately, due to feasibility issues we were unable to administer the WAB-R for the 3-month follow-ups; instead, we relied on subjective self-reports for the follow-up which are limited in reliability and may explain the failed detection of group differences in the secondary outcome measures.

### EEG

To better understand the mechanism and biomarkers of recovery, correlations between change in behavioral outcomes and change in EEG relative power and imaginary coherence were explored. As a reminder, both tDCS and sham groups received speech and language therapy, so interpretation of the mechanistic findings need to be mindful of the combined effects. While significant EEG – behavioral correlations in the tDCS group reflect both neuromodulation and speech therapy mediated recovery, significant correlations in the sham group represent primarily speech therapy mediated recovery with some placebo effects. Overall, we found that recovery in both groups relied heavily on the contralesional hemisphere, especially involving the T4 channel (right middle temporal). Recruitment of right homologous frontotemporal areas during language recovery is commonly observed in aphasia patients with left frontal stroke.^40,41^ We further found that language recovery for tDCS was mostly correlated with changes in the faster frequencies (e.g., alpha, beta) and for sham, the slower frequencies (e.g., theta, delta); see Supplementary Table 1 for both corrected and uncorrected correlation results. More specifically, for tDCS, increased relative alpha power at T4 significantly correlated with an overall language improvement as well as spontaneous speech, whereas for sham (placebo + SLT), decreased relative delta power at T4 was related to an overall language improvement.

Stroke injury typically leads to reduced higher frequencies and increased lower frequencies and recovery of higher frequencies has been associated with behavioral improvement.^9,10^ Specific to post-stroke aphasia recovery, higher alpha power at right frontal and temporal areas have been linked to better prognosis.^42^ These results are therefore in alignment with existing research. However, the sham group also revealed a positive correlation between change in relative delta power at F8 (right inferior frontal) and change in repetition. Comparing the correlation coefficient to that of tDCS yielded a significant group difference, suggesting a mechanistic difference between sham and tDCS in repetition recovery. Although for the sham group, repetition was one of the WAB-R scores that did not quite meet the cutoff, the trending improvement (*P* = 0.058) observed seems to be strongly related to an increase in delta at F8 (right inferior frontal) which was not observed after tDCS. Increase of delta frequencies during mental tasks have previously been associated with functional cortical inhibition and may modulate the activity of those networks that should be inactive to accomplish the task.^43^ Repetition primarily engages the motor network, with minimal involvement of Wernicke’s area. The completion of such task might be facilitated by inhibiting the right inferior frontal region in the case of a dysfunctional language network. A distinct mechanism seems to take place for patients receiving tDCS since a decreased beta coherence between T3 and T4 was a strong correlate of repetition improvement after treatment. Increased beta (and, particularly, beta mu) has been repeatedly associated with speech production in the literature.^44,45^ While we did not observe a significant increase in beta associated with repetition improvement, we found a trend of decreased beta at T3 related to improvement in another speech production task, name and word finding. Hypothetically, lesser involvement of the lesional site (as reflected by decreased beta at T3/left Broca) might facilitate the accomplishment of speech production tasks and be reflected by a decoupling in the amount of beta recorded at T3 and T4 after tDCS. As mentioned, this is only a hypothesis that should be more formally investigated in future studies.

To our knowledge, there are only two studies that have investigated changes in electrical brain activity following tDCS in post-stroke aphasia.^17,46^ Only one of these studies investigated tDCS of Broca’s area in a suboptimal sample (*n* = 6) using approximate entropy and did not find treatment related changes.^46^ Our findings provide insight into the potential mechanisms of recovery following tDCS, which appear to be distinct from the recovery associated with receiving SLT alone. Future studies should use tasks involving covert speech production, speech listening, and auditory discrimination during EEG to further understand the mechanisms underlying language improvements observed following tDCS.

### DTI

Extending and aligning with the EEG signatures of neuroplasticity in the right cortical regions, we also found that improvement in spontaneous speech after tDCS was associated with decreases in MD values of the left superior cerebellar peduncle (SCP), which was not found in the sham group. Mean diffusivity (MD), a proxy for white matter integrity, tends to increase after brain injury due to the loss of diffusion directionality.^47^ On the other hand, decreases in MD have been frequently found following intervention or training as a sign of neuroplasticity.^48–52^ MD decreases have also been associated with the brain-derived neurotrophic factor (BDNF) and may be indicative of long-term potentiation (LTP).^50^

The SCP is the main output pathway of the cerebellum and projects to the midbrain and thalamus with majority of its efferents decussating in the midbrain before passing to the contralateral thalamus^53^ and subsequent cortical regions. We speculate that neuroplastic changes in the left SCP may correspond to the compensatory recruitment of the right homologous cortical language areas associated with recovery, as observed in our EEG findings. The cerebellum, especially, the right posterolateral cerebellum, has been associated with a host of cognitive and language functions, beyond just motor functioning,^54–56^ where both functional and structural connectivity studies have identified connections between the right cerebellum and left fronto-temporo-parietal language association areas.^20,57–59^ While the right cerebellum coordinates with the left cortical language regions in the selection and production of words, in the case of left hemispheric damage associated with post-stroke aphasia, cerebellar activity has been found to switch hemispheres concurrent with recruitment of compensatory right homologous language regions.^60^ Because the cerebellum appears to be relatively spared in post-stroke aphasics, a few tDCS studies have recently focused on targeting the cerebellum as a non-lesioned gateway to the cortical language systems commonly affected by supratentorial stroke, especially for severe aphasic patients with extensive cortical lesions that may result in limited intact tissue for stimulation to be optimally effective. Cerebellar tDCS has produced improvements in various aspects of language performance in post-stroke aphasics, including naming,^61^ spelling,^62^ and verb generation.^63^ In healthy volunteers, cerebellar tDCS similarly resulted in better verb generation^64^ as well as verbal fluency.^65^ This is the first study to uncover a cerebellar white matter correlate of language recovery after tDCS treatment in post-stroke aphasia. Future studies should explore the language recovery mechanisms associated with cerebellar tDCS and identify patient characteristics that may predict a greater benefit from either cerebellar or Broca’s tDCS, to better personalize treatment and optimize outcome.

## Conclusion

In summary, the results of this study indicate that using anodal tDCS over Broca’s area in combination with speech and language therapy can effectively improve both expressive and receptive language abilities in individuals with chronic post-stroke aphasia. Our findings also provide new evidence that speech therapy paired with tDCS and sham/placebo may involve different recovery mechanisms, as assessed using EEG and DTI. These neuroimaging techniques provide valuable insights into the functional and structural changes occurring at both the cortical and subcortical levels after different interventions. The clinical implications of these findings include the potential to tailor neuromodulation strategies and enhance treatment outcomes for individuals with aphasia.

## Data Availability

The data underlying this article will be shared on reasonable request to the corresponding author.

## Non-standard Abbreviations and Acronyms

AQ: Aphasia Quotient
AVC: Auditory Verbal Comprehension
DTI: Diffusion Tensor Imaging
FA: Fractional Anisotropy
MD: Mean Diffusivity
NWF: Name and Word Finding
REP: Repetition
SLT: Speech and Language Therapy
SOM: Secondary Outcome Measures
SS: Spontaneous Speech
TSI: Time Since Injury
WAB-R: Western Aphasia Battery-Revised

